# Childhood to Adult Neurodevelopment in Gene-Expanded Huntington’s Disease (ChANGE-HD) study protocol: A Prospective Longitudinal Neurodevelopmental Study of Huntington’s Disease

**DOI:** 10.1101/2025.10.22.25338583

**Authors:** Mohit Neema, Nabil Halabi, Peggy C. Nopoulos, Michael Freedberg, the ChANGE-HD investigators, coordinators, and consultants

## Abstract

Although adult Huntington’s disease (HD) studies have significantly advanced our understanding of the course of degeneration, they may underrepresent critical neurodevelopmental aspects of the disease. Significant gaps remain in understanding how mutant huntingtin affects early neurodevelopment, its long-term impact, as well as potential implications for treatment outcomes. The Childhood to Adult Neurodevelopment in Gene-Expanded Huntington’s Disease (*ChANGE-HD;* NCT01951588) study aims to evaluate brain structure and function in premanifest, at-risk children and young adults, and explore HD’s developmental origins. Here, we introduce the ChANGE-HD study protocol, which will investigate and integrate the neurodevelopmental and neurodegenerative aspects of HD. The ChANGE-HD study is a prospective, multi-site observational trial with an accelerated longitudinal design. Four hundred participants aged 6-30 years who are at risk for HD will be recruited and asked to return for multiple visits (if possible). At each visit, cognitive, motor, behavioral, blood, and MRI data are collected. ChANGE-HD represents the first prospective multi-site study to systematically document brain structure and function during the premanifest phase of HD in children and young adults. Data collection is ongoing with first results anticipated in 2026-2027. The ChANGE-HD approach is likely to provide novel pathophysiological insights and guide the development of therapeutic strategies tailored to both the developmental and degenerative phases of the disease.

## INTRODUCTION

### Lessons from Kids-HD

Huntington’s disease (HD) is an autosomal-dominant neurodegenerative disorder of the central nervous system (CNS) resulting from an abnormal trinucleotide repeat expansion of cytosine-adenine-guanine (CAG) in the Huntingtin (*HTT*) gene [1]. In the general population, the number of CAG repeats encoded within exon 1 of *HTT* ranges from 10 to 35. Repeat lengths between 36 and 39 demonstrate variable penetrance, whereas repeats exceeding 39 are classified as mutant *HTT (mHTT)*, resulting in HD with full penetrance.

*HTT* is a highly conserved gene essential for nervous system development, neuronal stability, and survival. The gene is maintained across species [2], suggesting that CAG repeats within *HTT* have undergone positive selection, potentially driving species survival through increasing CNS complexity. Notably, humans exhibit the highest number of CAG repeats, underscoring their unique evolutionary significance to the development of higher-order cognitive tasks [3–5].

Traditionally, HD has been viewed as a neurodegenerative disorder primarily affecting the striatum, characterized by motor, cognitive, and behavioral dysfunctions [6]. The prevailing explanatory hypothesis attributes the disease to a toxic gain-of-function mutation within *mHTT* (i.e., the CAG repeat expansion), resulting in neuronal damage and cell death. However, emerging stem cell, animal, and human neuropathological evidence suggests that brain development changes are critical to HD’s pathoetiology [7–9], raising the hypothesis that HD is an unintended outcome of an adaptive evolutionary process supported by a more functional CNS [3]. Thus, *mHTT*, in addition to causing pathological effects in adulthood, may positively affect the course of neurodevelopment. Understanding the complex interplay between neurodevelopment and neurodegeneration could transform treatment approaches, particularly by identifying critical inflection points where neurodevelopment transitions into degeneration. Most show that the degenerative process in HD begins roughly 20-25 years prior to onset [10–12]. These findings underscore the need to understand the effects of HTT on development, particularly in children and young adults at risk for HD.

In 2009, the National Institute of Health (NIH) -funded Kids-HD study (ClinicalTrials.gov Identifier: NCT01860339) was launched at the University of Iowa, investigating the impact of *mHTT* on brain development in children at risk for HD [7]. For research purposes only, children aged 6-18 years-old at risk for HD (defined as having a parent/grandparent with HD) were genotyped and categorized as gene-expanded (i.e. GE, CAG >= 36) or gene-non-expanded (i.e. GNE, CAG < 36). The GNE children served as an excellent control group, given that being raised in an HD family can produce unique psychosocial strain not captured by children of non-HD families. The Kids-HD study modeled the progression of HD several decades before the predicted age of motor onset in the GE participants. Over the years, the Kids-HD study produced several key findings (Table 1). Some highlights include:

- **HTT impacts brain structure and function along the entire spectrum of repeats**: Higher repeats are associated with better cognitive skill and greater brain volume in GE and GNE individuals [13,14].
- **Methodological challenges of modeling age and CAG separately, versus years-to-onset model (which incorporates both):** The Kids-HD study employed an accelerated longitudinal design (ALD), the gold standard for evaluating brain development in children, which combines cross-sectional and longitudinal data to model age-related changes across a broad developmental window. This model showed not only interesting findings of accelerated striatal development, where GE subjects reached peak striatal volume years before the GNE group, but that these trajectories were profoundly affected by CAG repeat length [15]. We also found that every CAG repeat conferred a greater advantage in general cognitive ability [14]. This effect peaked at repeats of 43 and individuals with longer CAG repeats showed trajectories that were declining in function after age 15, likely representing changes due to the degenerative phase of the disease. The age-trajectory modeling results highlighted the profound consequences HTT and CAG repeat-length has on neurodevelopment. These findings recapitulate the well-established negative relationship between CAG repeat length and age of motor onset and require an updated model to take both factors, and their interactions, into account. Given the influence CAG repeats and age exert on longitudinal brain development, we began modeling the data in a Years-To-Onset (YTO) fashion to evaluate the time course of the disease from development to degeneration. The YTO model predicts the age of motor onset based on the participant’s current age and CAG repeat length. An example of the difference between an age-only trajectory and a YTO model is as follows: An 8-year-old with a CAG repeat length of 48 and a 22-year-old with a CAG length of 43 both have 26 YTO, despite being on opposite ends of the age trajectory model. When evaluating a biomarker of degeneration in Kids-HD (neurofilament light; “NfL”), we found that NfL levels are normal as far back as 40 YTO but rise around 20 YTO [16]. These findings support the notion that the YTO model can accurately characterize the developmental phase of the disease process (greater than 20 YTO).
- **The antagonistic pleiotropy theory of HTT and its implications for neurodevelopment:** In our most recent publication, we revealed that GE individuals from 50-20 YTO have larger cerebral volumes, more cortical surface area, and higher IQs compared to GNE subjects, but the opposite is true after 20 YTO [17]. Striatal and pallidal volumes remain comparable in volume to GNE until 20 YTO after which they dramatically decline in volume, consistent with early vulnerability of the basal nuclei. These findings challenge the traditional concept of HD as a uniquely neurodegenerative disorder and highlight the neurodevelopmental role of *mHTT.* Additionally, Kids-HD research uncovered potential early advantages of *mHTT* in GE children, such as larger brain development that supports enhanced cognitive development and reduced depression/anxiety. However, these children will indeed develop HD, highlighting the yoking of advantage with disadvantage. Our findings support the notion of antagonistic pleiotropy in HD, where the same neurodevelopmental differences that confer early advantage eventually lead to vulnerability and subsequent degeneration [18]. Collectively, they reshape our understanding of the evolutionary implications of *mHTT* and its potential role in brain development.

**Table 1.**
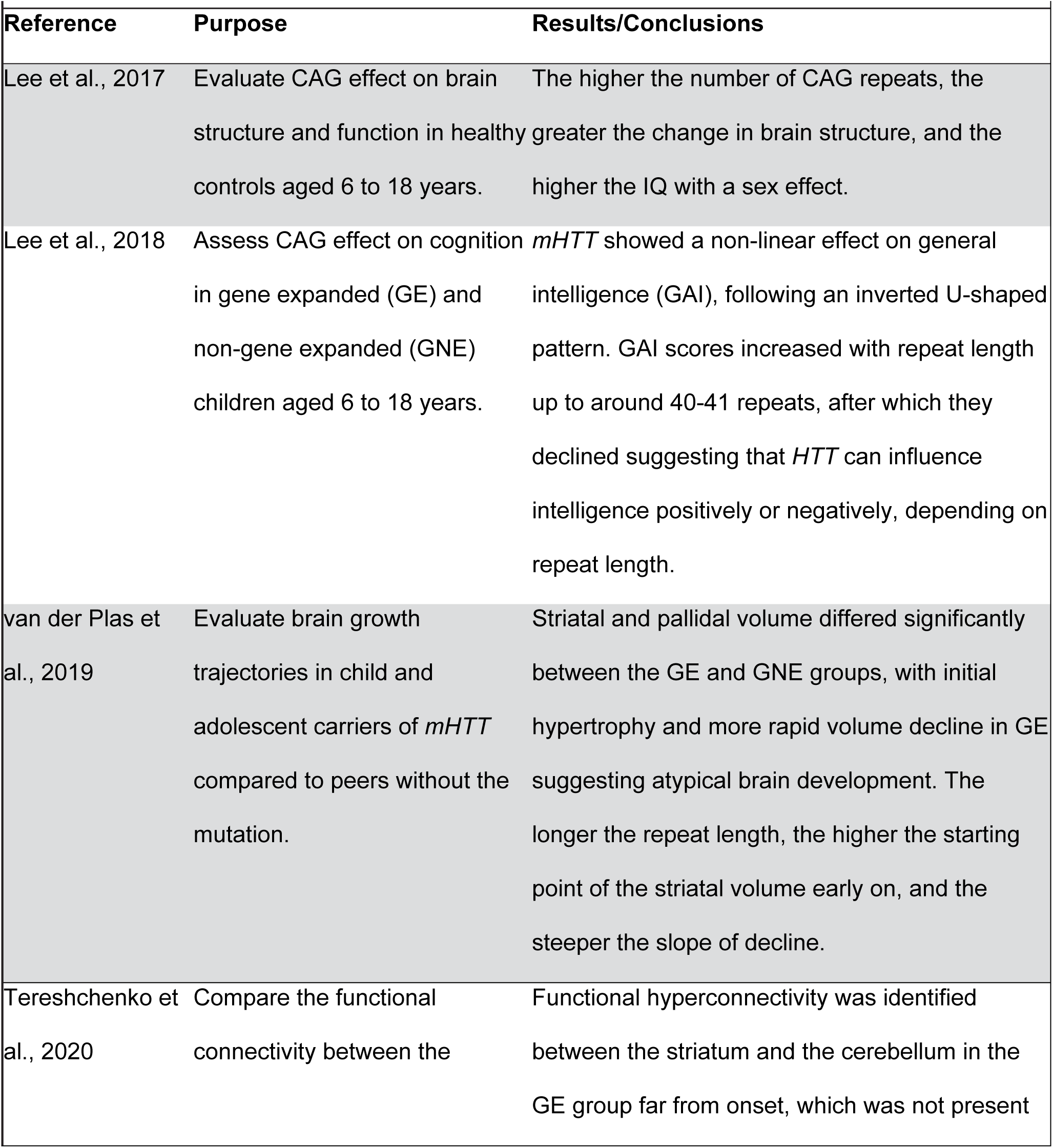
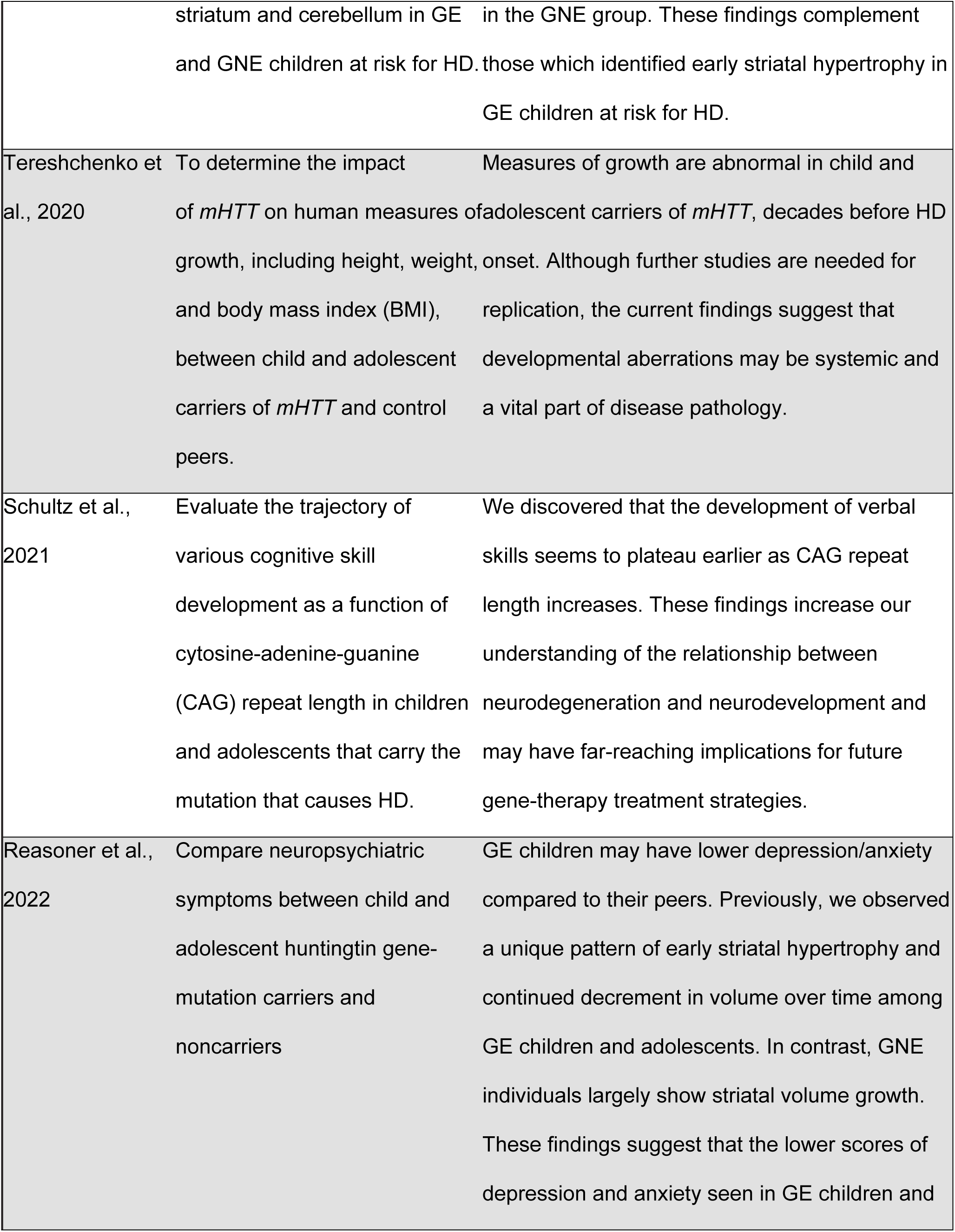
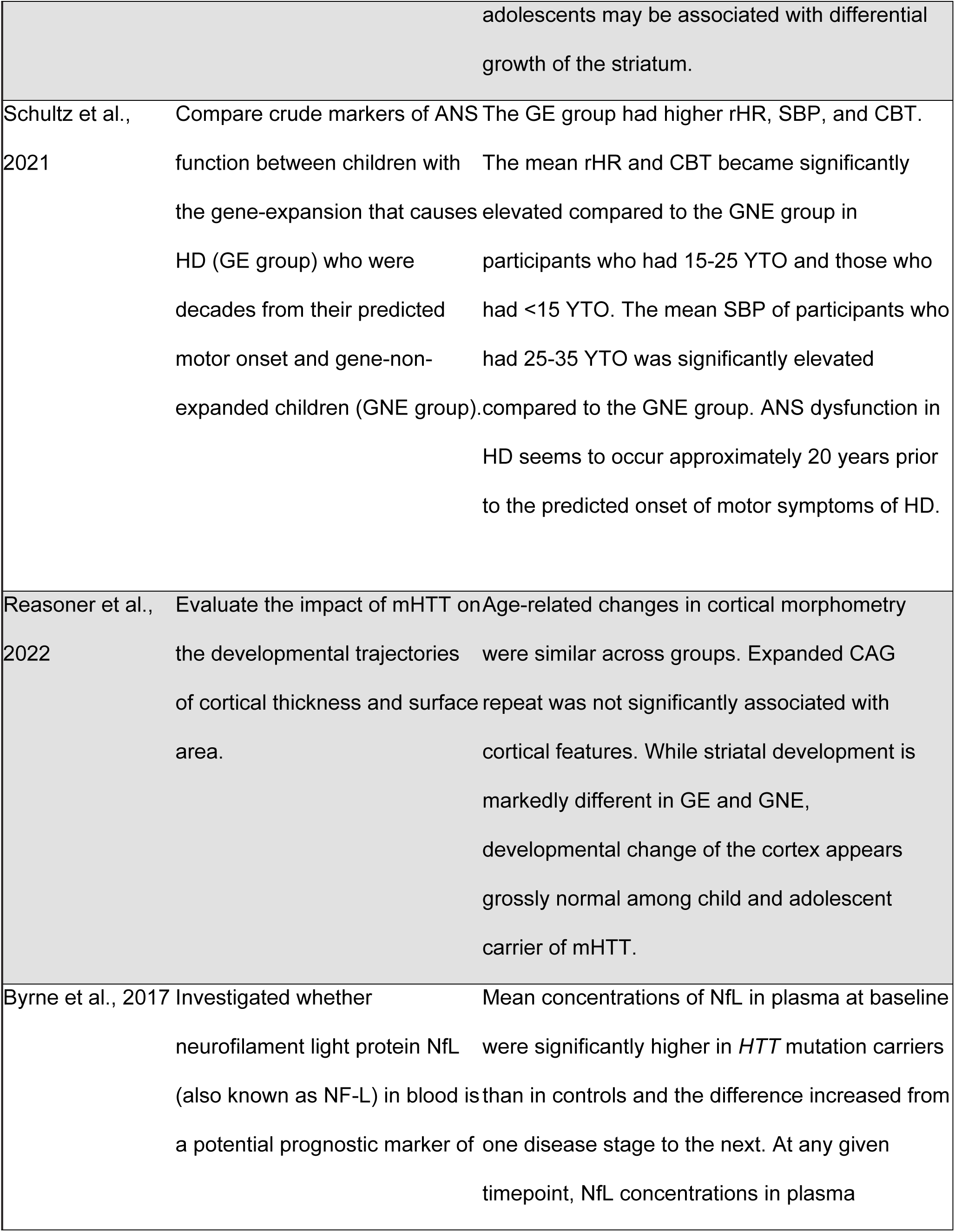
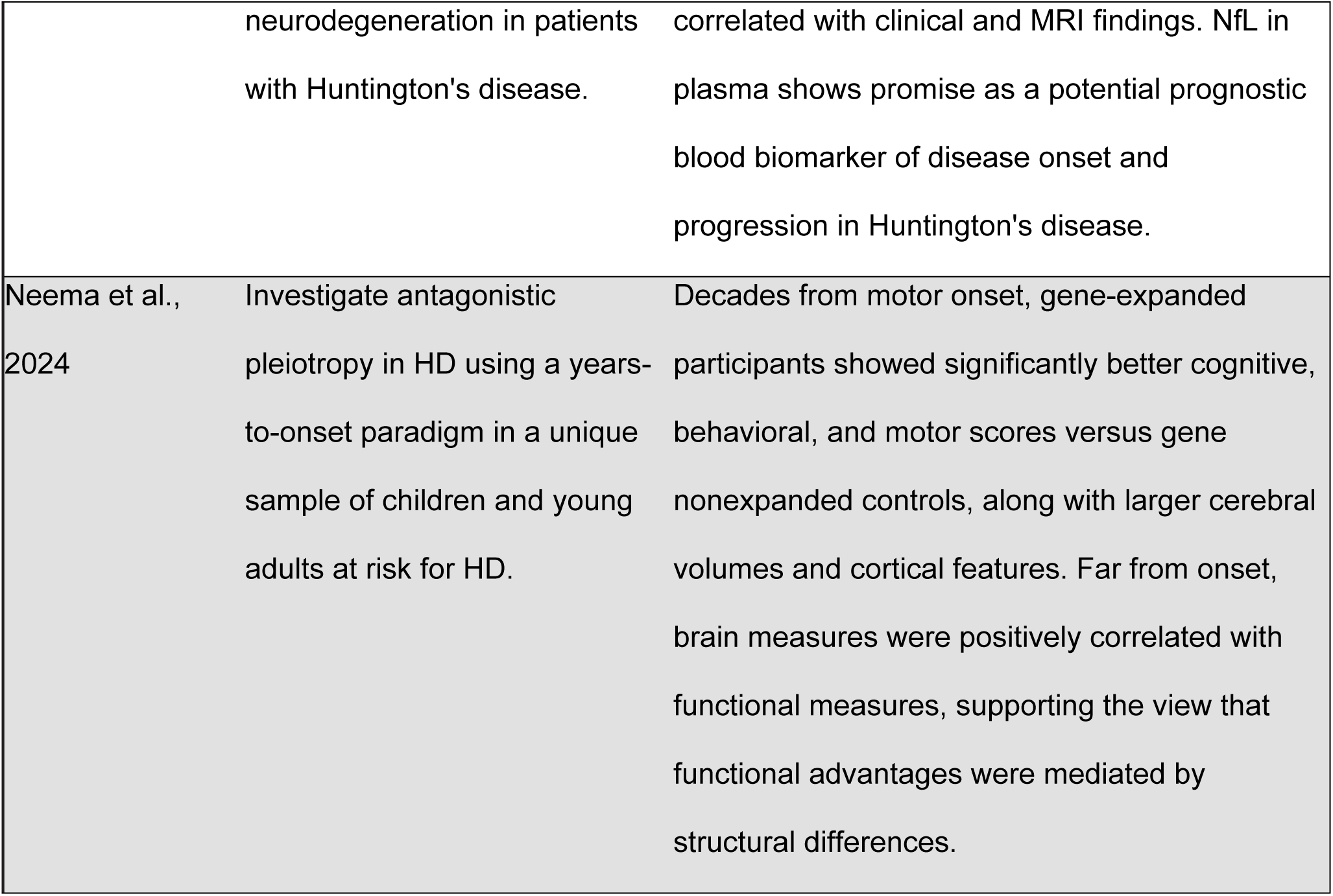
Summary of Kids-HD papers/findings.

### Beyond Kids-HD: Embracing a new era with ChANGE-HD

While the Kids-HD study provided novel insights, it also raised several critical questions that required further investigation. Kids-HD’s modest single-site sample (198 at-risk youth) produced valuable data but required a necessary expansion for greater reliability and broader application. This prompted the launch of the NIH-funded, multi-site ChANGE-HD study in 2019, which builds upon the Kids-HD findings and addresses the following key areas:

- **Expansion and Replication:** No single study can provide definitive conclusions. ChANGE-HD aims to replicate the Kids-HD findings using a larger sample size across multiple sites to ensure more robust, generalizable data.
- **CAG-specific effects:** Kids-HD identified CAG-specific effects in the GE group, where different repeat lengths produce distinct neurodevelopmental trajectories. To fully understand these effects, ChANGE-HD will recruit a larger cohort to enable a thorough analysis across a fuller range of repeats.
- **Wider age range amongst participants:** Kids-HD originally recruited participants from age 6 through age 18, but maturational changes extend into late 20s and early 30s [19]. Although YTO models address the strict need for sampling participants across a broad spectrum (as demonstrated by the example above), it is still beneficial to examine older participants who are approaching their date of motor onset to better model trajectories at low YTOs.
- **Evaluation of serum biomarkers of disease progression - Neurofilament light (NfL):** Serum NfL is a marker of neuronal damage in premanifest HD (among other neuronal diseases and disorders), with levels rising approximately 15–20 YTO and is strongly correlated with the age of motor onset. Although NfL is classically absent in the developmental phase and appears after the onset of subtle neurodegeneration, its use as a biomarker for HD remains unclear and requires further investigation. The CHANGE-HD study aims to clarify the trajectory of NfL in younger at-risk individuals and explore its relationship with clinical outcomes and MRI findings.

The goal of ChANGE-HD is to model development (versus degeneration) by focusing on individuals whose brain changes likely reflect neurodevelopment rather than disease progression. The broader objectives include:

1. **Assess brain structure and function using structural and functional MRIs to monitor brain development over time**. A specific focus is given to striatal circuitry, due to its centrality to HD and its prominence in earlier data.
2. **Evaluate the relationship between brain structure and functional outcomes by regressing neuroimaging findings (DTI, fMRI, structural MRI) against functional assessments (cognitive, behavioral and neurologic functioning).** Here, we aim to elucidate the precise effects of *mHTT* on cognition.
3. **Investigate the genotype-function relationship by examining the link between CAG repeat length and clinical (motor scores, functional assessments, cognitive evaluations) and biological (neuroimaging, biomarkers) measures**. This includes evaluation of neurofilament light (NfL) as an appropriate marker of neurodegeneration in HD.

The study objectives will be achieved by improving upon the Kids-HD infrastructure through multiple sites and YTO models. We expect the study to reveal the pre-manifest biomarkers of the disease and bridge the relationship between development and degeneration in HD.

## METHODS

### ETHICS (standard protocol approvals, consents, blinding, and safety)

Study approval and centralized oversight were provided by the WIRB-Copernicus Group (WCG; formerly WIRB; study number: 1269202; IRB Tracking Number: 20192908). WCG ensures compliance with federal regulations, ethical guidelines, local laws, participant safety, and study integrity. The study was conducted according to the principles expressed in the Declaration of Helsinki. Consent (or assent) procedures vary based on the age range of our participants.

Assent is a simplified version of the consent document presented in age-appropriate language:

- **Ages 6-7:** The assent form is verbally explained to minor participants, and they will choose whether to provide verbal assent. If verbal assent is provided, it is indicated on the physical consent document by the research team who explained the assent form to the minor participant. A parent or legal guardian reviews and chooses whether to provide full written consent form on the participant’s behalf.
- **Ages 8-11:** Literate participants have the assent form verbally explained to them, read, and decide whether to provide written assent. A parent or guardian then decides whether to provide written consent on their behalf.
- **Ages 12-17:** Both the participant and their parent or legal guardian review the full consent form with a research team member and whether to provide written consent.
- **Ages 18-30:** Adult participants review the full consent form with a research team member and decide whether to provide written consent on their own behalf.

Collecting predictive genetic data for HD in individuals below 18 years of age raises the risk of confidentiality and privacy breaches, the results of which can have highly negative psychological consequences on those individuals. Thus, a study of this kind warrants commensurate security measures. In accordance, all testing is performed in a double-blind manner and all research team members who interact with participants or their families remain blind to the participants’ genetic status. Those who have previously undergone genetic testing are asked not to disclose their results. Team members responsible for data analysis only have access to de-identified data and do not have any contact with participants or their families. At no point, including after the study’s completion, will any team member have access to both identifying information and genetic status. Genetic results are never shared with participants, their families, medical providers, or anyone with access to identifying information. Additionally, participants are free to withdraw from the study at any time. Data obtained prior to the time of withdrawal are retained, unless the participant or their caregiver specifically requests that all collected data be discarded. To verify that our protocols ascend to the appropriate level of security required to protect the records of all participants giving genetic samples in our study, the ChANGE-HD study protocols were submitted to several HD patient advocacy groups, including Help4HD International, the Huntington’s Disease Society of America (HDSA), and the Huntington’s Disease Youth Organization (HDYO). All groups have reviewed and endorsed our security measures.

### DESIGN

The ChANGE-HD study employs an accelerated longitudinal design (ALD), combining cross-sectional and longitudinal approaches by recruiting participants across the desired age range. Since the ALD allows data collection at different time points across participants’ ages, follow-up testing is requested but is not a strict requirement for participation. The clinical and imaging protocols are modeled after the ‘Adolescent Brain Cognitive Development’ (ABCD) study, providing a well-established framework for cognitive, behavioral, and MRI assessments in a longitudinal multi-site setting.^18^ Furthermore, the protocol has been modified such that all measures are standardized for comparison across the range of 6 to 30 years-of-age.

### SITES

At the study’s commencement in 2019, there were five sites: the University of Iowa, the University of California, Davis, the University of Texas, Houston, the Children’s Hospital of Pennsylvania, and Columbia University. The four additional sites were carefully selected for their location, patient population, and strong HD research infrastructure. Vanderbilt University was added in 2022.

### STUDY PERIOD AND RECRUITMENT GOALS

ChANGE-HD was originally planned as a 5-year study, aiming to complete 1,500 assessments of 400 participants by the end of the fifth year. However, the COVID-19 pandemic delayed study progress by causing significant disruptions in recruitment and scheduling across the five original study sites and requiring us to update our timelines (*cf.* Table 2). As a result, the assessment period has been extended to approximately seven years. Despite the disruptions of COVID-19, our most recent power analysis shows that we are still on track to achieve our study aims.

**Table 2.**
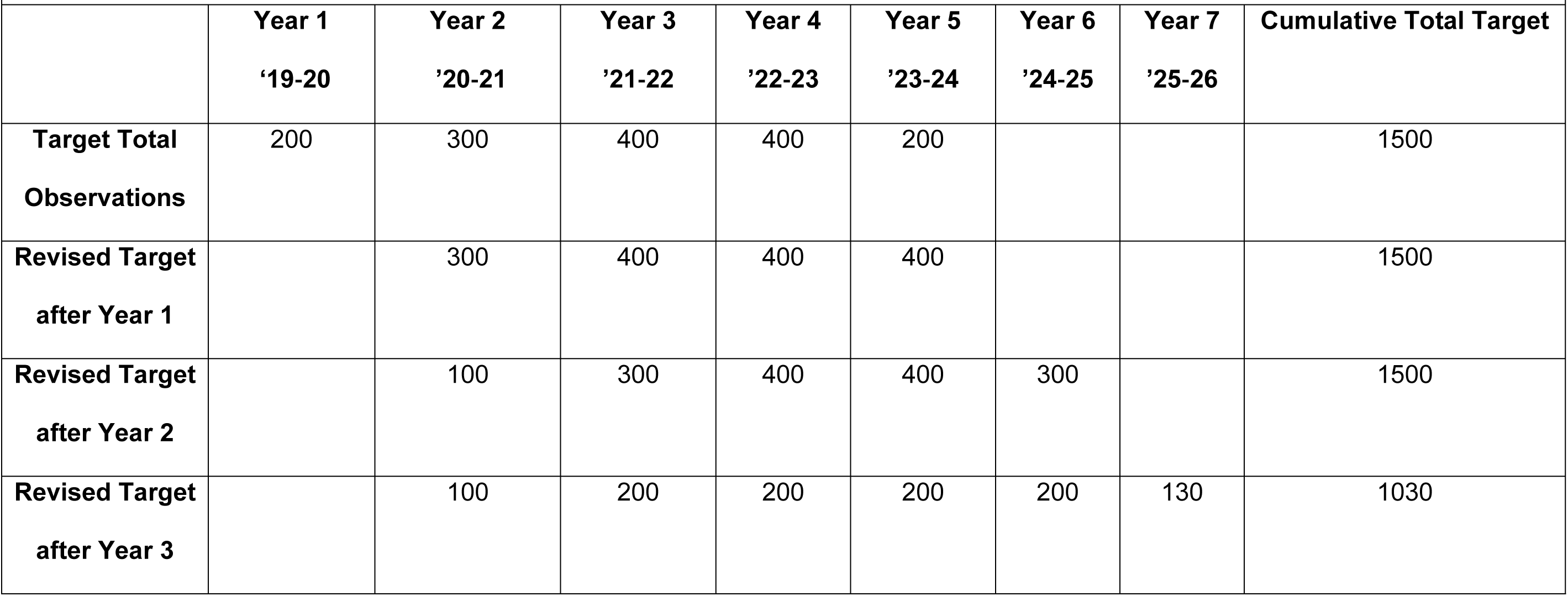
Study timeline and recruitment target in study visits.

Our study aims were to evaluate the effects of gene status on developmental trajectories of brain structure (volumes) and brain function (cognition, behavior, motor skill) and our findings from Kids-HD, showing greater cerebral cortex volume and cognitive ability from 50-30 years from motor symptom onset in GE compared to GNE participants [17]. For the key significant findings in that paper, we should have at least 90% power to replicate the findings at p = .01. This conclusion is based on the following analysis: Our final sample size projection is 1,030 observations from at least 450 separate participants. Assuming 40% of participants are gene-expanded (as in the previous study), we can assume at least 400 visits from 180 unique genetically affected participants. The previous study was based on 136 visits from 79 gene-expanded participants. A conservative estimate of the relative information currently collected compared to the previous study is therefore obtained by the ratio of unique participants, which is 2.28. (The ratio of total visits is higher.) The findings from our Kids-HD study documented in Table 3 can be translated to non-centrality parameters (NCP) for those findings, which are then readily translatable to statistical power. Assuming those findings are reproducable, expected non-centrality parameters for the current study will be at least 2.28 times larger. We compare these projected NCP values (Table 3) to critical NCP thresholds for the above-claimed power (15.1, 19.8, and 22.9 for 1, 3, and 5 numerator df, respectively). Thus, our adjusted recruitment target is 450 participants and 1,030 observations over a 7-year timeline across six sites. Currently, data collection is ongoing with 457 participants recruited and 914 observations recorded. Participant recruitment and data collection are expected to end in March of 2026, with first results expected to be published in 2026-2027.

**Table 3.** Findings from Neema et al. (2024). NCP is calculated from an F statistic by NCP = (F-(2F/ddf)-1)*ndf. NCP = non-centrality parameter, ddf= denominator degrees-of-freedom, ndf = numerator degrees of freedom.

| Measures | N | Num DF | Den DF | F | p | Observed NCP | Projected (NCP*2.28) |
| --- | --- | --- | --- | --- | --- | --- | --- |
| General Ability Index | 136 | 1 | 87.66 | 13.5 | <0.001 | 12.19 | 29.44 |
| Behavior composite | 135 | 2 | 120.27 | 10.91 | <0.0001 | 19.46 | 47.34 |
| PANESS | 128 | 1 | 46.12 | 8.92 | 0.005 | 7.53 | 18.93 |
| Whole brain | 117 | 3 | 62.63 | 7.79 | <0.001 | 19.62 | 49.49 |
| Cerebrum | 117 | 3 | 62.55 | 7.8 | <0.001 | 19.65 | 49.56 |
| Cortical gray matter | 117 | 1 | 99.9 | 12.22 | <0.001 | 10.98 | 26.59 |
| Cortical surface area | 117 | 1 | 95.22 | 12.17 | <0.001 | 10.91 | 26.47 |
| Cortical thickness | 117 | 1 | 54.67 | 2.95 | 0.091 | 1.84 | 5.61 |
| Cortical folding index | 117 | 3 | 82.04 | 5.54 | 0.002 | 13.21 | 34.46 |
| Cortical curvature index | 117 | 1 | 62.27 | 10.49 | 0.002 | 9.15 | 22.56 |
| Cerebral white matter | 117 | 3 | 51.46 | 9.84 | <0.0001 | 25.37 | 63.10 |
| Cerebellum | 117 | 2 | 74.95 | 9.67 | <0.001 | 16.82 | 41.54 |
| Striatum | 117 | 5 | 102.37 | 13.98 | <0.0001 | 63.53 | 152.87 |
| Globus pallidus | 117 | 5 | 80.15 | 16.22 | <0.0001 | 74.08 | 177.72 |
| Thalamus | 117 | 1 | 102.87 | 7.47 | 0.007 | 6.32 | 15.87 |

### PARTICIPANTS

At baseline, approximately 400 total participants will be enrolled across all sites. Although annual visits are not required for an ALD design, retention is best with close follow-up, and subjects are asked to return annually. We aim to recruit an even distribution of subjects across age groups, with 200 participants in the range of 6-18 years-old and the remaining participants in the range of 19-30 years-old. To participate, participants must:

- Be between 6-30 years old.
- Have a biological parent or grandparent who has been diagnosed with HD, indicating that the participant is at risk for HD.
- Demonstrate an age-appropriate understanding of HD and its risks. If the parents indicate that their child is unaware, we will exclude the participant until an age-appropriate discussion has occurred.
- Not have a history of medical diagnoses that could potentially confound the study results (e.g., major head trauma, seizures, tumors, or any other major medical illness requiring ongoing care). If an aspect of their medical history obscures their symptoms as being directly attributable to HD (e.g., a concomitant congenital disorder) they are excluded from participation.
- Be fluent in English.
- Not have experienced Juvenile-onset HD (JOHD).
- Not have manifested overt motor features of HD. This is ascertained by reporting from parents (or participants themselves, if they are over 18) during a screening visit. If the participant or the parent reports that they are currently manifesting any motor symptom of HD, the participant is referred to a child or adult neurologist for clinical evaluation, respectively.
- Be free of MRI contraindications. Participants of childbearing potential must undergo a urine pregnancy test before their MRI. If positive, they are excluded from the MRI but may still complete other assessments.

Participant recruitment is conducted across the United States. Potential participants along with their parents or grandparents are identified through local hospital or clinic records and HD registries. Recruitment platforms such as *Research Match*, *HD Trial Finder*, and *ClinicalTrials.gov* are used to share study information with the community. Additionally, study-dedicated social media channels and websites are maintained to disseminate updates. The study is also advertised through events, newsletters, social media, and websites of national HD organizations, including the HDSA, its state chapters, Help4HD International, National Youth Alliance (NYA) of HDSA, HDYO, and the Huntington Study Group (HSG).

### DATA COLLECTION AND ASSESSMENTS

The following assessments are completed at each study visit (see *Table 4* and *Supplementary table 2* for more details):

**Table 4.** Procedures performed at each ChANGE-HD visit.

| **Table 4.** Procedures performed at each ChANGE-HD visit. |  |  |  |  |
| --- | --- | --- | --- | --- |
|  | **Baseline**<br>**Visit 1** | **12 Month**<br>**Visit 2** | **24 Month**<br>**Visit 3** | **36 Month**<br>**Visit 4** |
| **Screening** | • | • | • | • |
| **Consent** | • | • | • | • |
| **Demographics \*** | • | Review and update if changed |  |  |
| **Vitals \*** | • | • | • | • |
| **Urine pregnancy screen \*** | • | • | • | • |
| **Blood draw \*** | • | • | • | • |
| **Blood processing \*** | • | • | • | • |
| **Saliva \*\*** | • |  |  |  |
| **Birth history \*** | • | Review and update if not collected previously |  |  |
| **Medical history \*** | • | Review and update if changed |  |  |
| **Family history \*** | • | Review and update if changed |  |  |
| **Medications \*** | • | Review and update if changed |  |  |
| **Behavioral protocol (Self and Proxy)** | • | • | • | • |
| **Cognitive protocol** | • | • | • | • |
| **Motor protocol** | • | • | • | • |
| **MRI** | • | • | • | • |
| * All females of child-bearing potential will be tested for pregnancy; ** Only completed at Visit 1 if blood draw is rejected; Details of all these measures are provided in the supplemental table. |  |  |  |  |

#### 1) BIOLOGICAL SAMPLES

Blood samples are collected via venipuncture at each visit using kits provided by Biospecimen Exchange for Neurological Disorders (BioSEND), an NIH-funded biorepository at Indiana University. If the blood draw is unsuccessful, a salivary sample is collected instead. After collection, samples are shipped to BioSEND for central storage and analysis. BioSEND will first perform genetic testing to determine the CAG repeat length within exon 1 of the *HTT* gene on chromosome 4. Additionally, plasma NfL concentrations will be measured using the *NF-Light® assay* with the *Simoa HD-1 analyzer* in collaboration with the Queen Square Institute of Neurology, University College London, London, United Kingdom.

#### 2) FUNCTIONAL MEASURES

As mentioned previously, the clinical assessment protocol is modeled after the ABCD Study. *Table 5* details the full assessment battery, including measures of cognition, behavioral health, environment & health, and motor function.

**Table 5.** ChANGE-HD assessments.

| Table 5. ChANGE-HD assessments |  |  |
| --- | --- | --- |
| Domain | Test Name | Construct |
| Cognitive | NIH TB - Picture Vocabulary | Language |
|  | NIH TB - Oral Reading Recognition | Language |
|  | NIH TB - Dimensional Change Task | Executive function |
|  | NIH TB - Flanker Inhibitory Control | Attention |
|  | NIH TB - List Sorting Working Memory | Working memory |
|  | NIH TB - Picture Sequence Memory | Episodic memory |
|  | NIH TB - Pattern Comparison Processing Test | Processing speed |
|  | NIH TB - Rey Auditory Verbal Learning Task | Verbal memory |
|  | Controlled Oral Word Association | Verbal fluency |
| **Motor** | Unified Huntington's Disease Rating Scale (UHDRS) | Motor |
|  | NIH TB - 9-hole Pegboard Task | Dexterity |
|  | Q-Motor | Motor |
| **Behavioral** | Achenbach System of Empirically Based Assessment | Adaptive and maladaptive functioning |
|  | UPPS-P Impulsive Behavior Scale | Impulsivity |
|  | Behavioral Inhibition/Behavioral Approach System | Inhibition/Reward seeking |
|  | Child and Youth Resiliency Measure & Adult Resiliency Measure, 5-pt scale versions | Resilience |
|  | Sleep Disturbance Scale for Children | Sleep |
|  | Sports and Activities Involvement Questionnaire | Involvement in sports, music, hobbies |
|  | Ohio State TBI Screen-Short | Traumatic brain injury of youth |
|  | Substance Abuse and Substance Use Form (SASUF) | Substance use/abuse |
| **Environment and Health** | Neuro-QoL short forms:<br>Positive Affect and Well-being (Adult)<br>Emotional and Behavioral Dyscontrol (Adult),<br>Anger (Pediatric) | Mental health |
|  | Neuro-QoL short forms:<br>Ability to Participate in Social Roles and Activities (Adult) | Social health |
|  | Satisfaction with Social Roles and Activities<br>(Adult)<br>Social Relationship - Interaction with Peers<br>(Pediatric) |  |

In the Kids-HD study, cognitive evaluation relied heavily on the Wechsler scales. However, in alignment with new NIH guidelines seeking to improve the inter-rater reliability of multi-site cognitive outcomes, a protocol change was implemented to use the NIH Toolbox (Version 2) [20]. The NIH Toolbox is an iPad-based assessment tool designed to obtain comprehensive metrics from a single visit in the following domains: language, verbal fluency, executive function, attention, working memory, episodic memory, processing speed, and visuospatial intelligence. It offers several advantages, including being digital, standardized, and comprehensive, reducing variability in test results, improving the precision of cognitive measurements, and enabling comparisons across diverse ages and sites.

Motor functions are assessed using the Unified Huntington’s Disease Rating Scale (UHDRS) motor battery, the Nine Hole Peg Test, and the Quantitative Motor (Q-Motor) assessment. The Q-Motor battery assesses tapping speed and regularity of movements produced by index fingers, hands, and feet. It also quantifies grasping, lifting, and involuntary choreiform movements. Altogether, it provides a comprehensive and precise evaluation of motor function that has been validated in children [21].

We measure behavioral metrics including participants’ adaptive and maladaptive functions, impulsivity, inhibition/reward seeking, and psychological resiliency using the Achenbach System of Empirically Based Assessment, the UPPS-P Impulsive Behavior Scale, the Behavioral Approach System, and the Adult/Child-Youth Resilience Measure, respectively. Finally, we also record participants’ physical, mental, and social well-being, sleep, the presence of sleep disorders, physical activities, history of traumatic brain injury, and substance use and abuse using Neuro-QoL [22], the Sleep Disturbance Scale for Children [23], the Sports and Activities Involvement Questionnaire, the Ohio State TBI Screen (short version) and a substance abuse and use form (SASUF, made in-house), respectively.

#### 3) MAGNETIC RESONANCE IMAGING

The MRI protocol is designed to collect high resolution structural and functional MRI data within the time span of a typical research MR exam. The scanning protocol is adapted from the harmonized ABCD study [24]. Specifically, we use 3 Tesla research scanners to collect T1-, T2-, and diffusion-weighted images, and resting-state functional MR brain images (see *Tables 6-9* for sequence parameter details). These sequences have been adapted and optimized for all 6 sites across Philips, GE, and Siemens MR scanner platforms.

**Table 6.** T1-weighted Scans. All sites used an acceleration factor of 2, a flip angle of 8 degrees, resolution was 1 mm isotropic, and phase partial Fourier was off. FOV = field of view, TR = repetition time, TE = echo time, TI = inversion time

| Site | Vendor | Model | Matrix | FOV | % FOV Phase | TR(ms) | TE(ms) | TI(ms) | Duration |
| --- | --- | --- | --- | --- | --- | --- | --- | --- | --- |
| Iowa | GE | 750W | 256x256x208 | 256x256 | 100% | 3000 | 2.0 | 1060 | 7:12 |
| CHOP | Siemens | Prisma | 256x256x176 | 256x256 | 100% | 2500 | 3.7 | 1060 | 7:26 |
| Columbia | GE | 750W | 256x256x208 | 256x256 | 100% | 7684 | 3.0 | 1060 | 7:20 |
| Houston | Philips | Ingenia | 256x256x225 | 256x256 | 93.75% | 6.658 | 2.9 | 1060 | 5:52 |
| Davis | Siemens | TrioTim | 256x256x176 | 256x256 | 100% | 2300 | 3 | 1060 | 5:48 |
| VUMC | Phillips | Ingenia | 256x256x225 | 256x240 | 93.75% | 6900 | 3.2 | 1060 | 5:52 |

**Table 7.** T2-weighted Scans. For all sites, resolution was 1 mm isotropic, field of view was 256×256, the field of view percentage was 100%, and phase partial Fourier was off. TR = repetition time, TE = echo time.

| Site | Vendor | Model | Matrix | TR (ms) | TE (ms) | Flip Angle (deg) | Acc. Factor | Duration |
| --- | --- | --- | --- | --- | --- | --- | --- | --- |
| Iowa | GE | 750W | 256x256x208 | 3200 | 60 | var | 2x | 6:28 |
| CHOP | Siemens | Prisma | 256x256x176 | 3200 | 564 | 120 | 2x | 4:52 |
| Columbia | GE | 750W | 256x256x204 | 3202 | 55.7 | 90 | 2x | 4:58 |
| Houston | Scanner | Ingenia | 256x256x256 | 2500 | 251.7 | 90 | 2.5x | 2:53 |
| Davis | Siemens | TrioTim | 256x256x176 | 3200 | 565 | T2var | 2x | 4:42 |
| VUMC | Philips | Ingenia | 256x256x256 | 2500 | 268 | 90 | 2.5x | 2:50 |

**Table 8.** Resting-state fMRI Scans. For all scans, the flip angle was 52 degrees, the TE was 30 ms, the field of view was 216×216, the field of view percentage was 100%, and the phase partial Fourier was off. The ABCD-Philips half scan factor (Houston and VUMC) was 0.9. TR = repetition time.

| Site | Scanner Vendor | Model | Matrix | Resolution (mm) | TR (ms) | Acc. Factor | Duration |
| --- | --- | --- | --- | --- | --- | --- | --- |
| Iowa | GE | 750W | 90x90x60 | 2.4x2.4x2.4 | 1100 | 4 | 5:00x4 |
| CHOP | Siemens | Prisma | 90x90x60 | 2.4x2.4x2.4 | 800 | 6 | 5:00x4 |
| Columbia | GE | 750W | 90x90x60 | 2.4x2.4x2.4 | 1365 | - | 5:00x4 |
| Houston | Philips | Ingenia | 96x96x60 | 2.25x2.25x2.4 | 800 | 6 | 5:00x4 |
| Davis | Siemens | Tim Trio | 86x86x60 | 2.5x2.5 | 800 | 6 | 4:08x4 |
| VUMC | Phillips | Ingenia | 96x96x60 | 2.25x2.25x2.4 | 800 | 6 | 5:15x4 |

**Table 9.** Diffusion-weighted Scans. For all scans the field of view percentage was 100%, and parallel imaging was off. For Philips scanners, phase partial Fourier was half of scan factor, FOV = field of view. TR = repetition time, TE = echo time.

| Site | Vendor | Model | Matrix | FOV | Resolution<br>(mm) | TR<br>(ms) | TE<br>(ms) | Flip Angle<br>(deg) | Acc.<br>Factor | b-values<br>(directions) | Phase<br>Partial<br>Fourier | Duration |
| --- | --- | --- | --- | --- | --- | --- | --- | --- | --- | --- | --- | --- |
| Iowa | GE | 750W | 140x140x81 | 240x240 | 1.7x1.7x1.7 | 9000 | 102 | 90 | 3 | 500(6)<br>1000(15)<br>2000(15)<br>3000(60) | 5.5/8 | 14:36 |
| CHOP | Siemens | Prisma | 140x140x81 | 240x240 | 1.7x1.7x1.7 | 4100 | 88 | 88 | 3 | 500 (6)<br>1000 (20)<br>2000 (20)<br>3000 (64) | 6/8 | 13:57 |
| **Columbia** | GE | 750W | 140x140x81 | 240x240 | 1.7x1.7x1.7 | 10000 | 101.7 | 90 | 2 | 3000 (96) |  | 17:30 |
| **Houston** | Philips | Ingenia | 144x144x80 | 240x240 | 1.7x1.7x1.7 | 6500 | 96 | 78 | 3 | 500 (6)<br>1000 (15)<br>2000 (15)<br>3000 (60) | 0.603 | 11:07 |
| **Davis** | Siemens | TrioTim | 122x122x78 | 241x241 | 2x2x2 | 4170 | 103 | 90 | 3 | 500(6)<br>1000(15)<br>2000(60) | 6/8 | 7:05 |
| **VUMC** | Phillips | Ingenia | 144x144x80 | 240x240 | 1.7x1.7x1.7 | 6500 | 96 | 78 | 4 | 500 (6)<br>1000 (15)<br>2000 (15)<br>3000 (60) | 0.603 | 11:11 |

#### 4) DEMOGRAPHICS AND HISTORY

Once cognitive, behavioral, motor, and environmental assessments have been administered, demographic and medical historical data are recorded. The parent or adult participant fills out demographic, family, and medical history forms in addition to completing a medication log. Finally, we record anthropometrics, including height, weight, head circumference, blood pressure, resting heart rate, and body temperature.

#### 5) DATA COLLECTION AND MANAGEMENT

REDCap (*Research Electronic Data Capture*) is an encrypted, HIPAA-compliant web-based application used in ChANGE-HD for data entry and storage, eliminating the need for double data entry and verification. Internal audits of de-identified data are performed regularly to ensure accurate data entry and monitor interrater reliability.

### DATA ANALYSIS

#### MRI Processing and analysis

Images from all sites are collected and imported into Flywheel, an infrastructure platform that allows researchers to manage, view, and analyze imaging data. Flywheel “gears” are containerized applications that automate reproducible analysis pipelines across all sites. Flywheel facilitates image sharing and automates analysis of volumetric, diffusion-based, and resting-state MR data.

- **Structural/volumetric analysis:** We use *volBrain’s AssemblyNetZ* [25] and *FreeSurfer* [26] on anatomical T1-weighted volumes to obtain cortical volume, surface area, and thickness, and subcortical grey region volumes. Our primary volume measures include the basal nuclei and sensorimotor cortex.
- **Structural connectivity/Diffusion MR analysis**: We use diffusion MR and diffusion tensor imaging (DTI) to measure water molecule diffusion, allowing us to infer the integrity of white matter microstructure and the developmental maturity of brain tracts. High resolution, whole brain diffusion MR is acquired with multiple diffusion sensitivities (“b-values”) and phase encoding directions, which allow us to correct geometric distortions in echo planar volumes. Diffusion MR preprocessing is performed with the QSIPrep Flywheel gear QSIPrep [27]. QSIPrep is an automated pipeline that performs denoising, distortion correction, motion correction, and co-registration to T1 volumes and atlas spaces. Quality assurance (QA) reports on motion, noise, and image quality are generated at the subject level for review. The output of QSIPrep is then passed to two further diffusion MR analysis gears that measure diffusion parameters such as fractional anisotropy in regions of interest (ROIs) and tractography based structural connectome metrics.
- **Resting-state functional connectivity MRI analysis**: During resting state scans, participants are asked to remain awake and keep their eyes open while breathing and blinking normally and blood oxygen level dependent (BOLD) signal is measured. After signal is collected, the functional organization of the brain can be inferred by regressing the BOLD time series of multiple ROIs and network against each other, where positive correlations infer synchronous communication, negative values infer asynchronous communication, and near-zero or zero values infer little or no communication. Findings from resting state data are robust and reliable, revealing consistent functional networks [28] and strong intra-subject reliability [29]. We will use data from resting-state scans to analyze and understand functional neurodevelopment in GE subjects and how it compares to GNE subjects. Resting-state fMRI preprocessing is performed with fMRIPrep [30], which provides denoising, motion correction, susceptibility distortion correction, and alignment to each subject’s T1 anatomical image and atlas space. The preprocessed data are then analyzed using XCP-D [31], which performs nuisance regression and temporal filtering and produces brain parcellated connectivity matrices. These matrices are generated using a cortical and subcortical atlas matched to the diffusion MR atlas, enabling direct comparison between structural and functional connectivity. For the ChANGE-HD study, the primary measures are pairwise functional connectivity values and network-level metrics related to striatal-cortical circuits.

#### Statistical analysis using YTO Models

ChANGE-HD employs an ALD, which is widely regarded as the gold standard for assessing developmental changes in brain development in children [32]. Instead of a traditional longitudinal design with two necessary timepoints separated by a specified interval, all visits across all participants are combined to create the largest possible pool of observations. This complementation across hundreds of participants allows for data to be obtained across a wide range of YTO. These data are then analyzed using linear mixed-effects models with random-effect intercepts accounting for inter- and intra-subject variability.

As mentioned above, one of the take-aways from Kids-HD was the insufficiency of using age-based or CAG-based models alone. Because the number of CAG-repeats influences age-related disease trajectories, a model relying solely on one parameter or the other will underdetermine the predicted age of motor onset. Furthermore, only 73% of the variance in age at motor onset can be attributed to CAG repeat lengths alone, indicating that other factors must be considered for accurate modeling. Inspired by this problem, Langbehn et al. (2004) designed a parametric survival model which incorporates both CAG and age effects to accurately produce a 95% confidence interval for a given patient’s predicted age of motor onset [33]. This survival model has the added benefit of being constructed using patients who have achieved motor onset *in addition* to asymptomatic GNE individuals, thus empowering the model with data from patients across the disease spectrum. Moreover, the ‘time-to-event’ modelling is applied across a full range of CAG lengths and ages, thus capturing a wide array of CAG/age combinations.

The general methodology of constructing the YTO models is as follows. First, the observed distribution of ages at onset from a large international sample of manifest HD patients was fit to a logistic distribution (which we found modeled the data significantly better than the Cox survival model). Next, we identified mathematical functions which best explain the relationship between CAG lengths and the mean/dispersion of age at onset for GE and GNE participants. Briefly, smooth curves are created by fitting “splines” (piece-wise polynomials) to the data and bends in the age trajectory are connected at the intersection of splines, called “knots.” Unique parameters for age at onset are then identified at each CAG length. The final model combines the best fit parametric distribution with the appropriate mathematical functions to create the final survival model. In general, the resultant model adopts the form of a conditional probability: Given that a presymptomatic individual has not yet achieved motor onset by age *x*, how likely is it that they will achieve onset in each preceding year? It is important to note that this model is optimized for individuals with CAG repeats between 41 and 56. The results may be biased for repeats below 41 due to heterogeneity in the distribution of observations for individuals with a CAG < 41 across the different study sites. Equation 1 represents the detailed function.

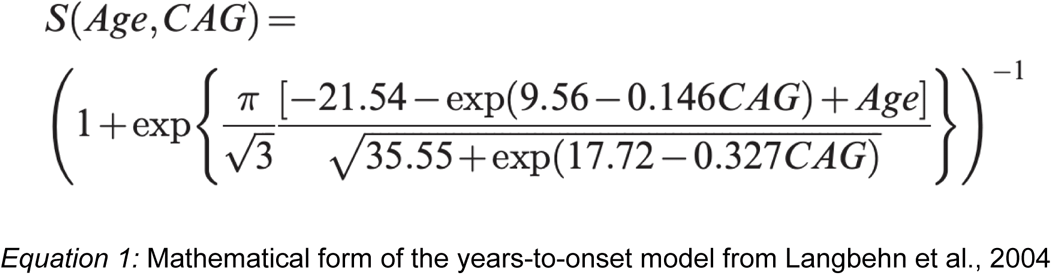

## THE JUVENILE-ONSET HD (JOHD) STUDY

Although most HD patients typically manifest obvious motor signs between the ages of 40 and 50 (known as Adult-Onset HD, AOHD), a much smaller percentage of patients are diagnosed with motor features before the age of 21 (classified as Juvenile-Onset HD, JOHD) [34]. Patients with JOHD experience the same triad of cognitive, behavioral, and motor symptoms as those with AOHD, albeit with distinct phenotypes. Specifically, JOHD patients tend to exhibit less hyperkinesia and more hypokinesia compared to AOHD. However, due to its rarity there is a lack of comprehensive data on the neurobiology and progression of JOHD.

While large-scale observational studies of AOHD have significantly advanced our understanding of HD, JOHD patients have been largely excluded from these efforts [35]. As such, large-scale longitudinal studies in JOHD are needed to deepen our understanding of the pathophysiology, as well as therapeutic strategies. When the Kids-HD study began at the University of Iowa in 2009, children at risk for HD who were already manifesting motor signs were followed as well. This was called the Kids-JOHD study (which ran in parallel to the Kids-HD study), whose primary objective was to deepen our understanding of the course and progression of JOHD. Cross-sectional analyses showed that patients with JOHD have significant subcortical degeneration shortly after diagnosis, along with reduced general measures of growth (such as weight and BMI) compared to GNE controls [36–38]. Furthermore, the longitudinal trajectory of disease severity is markedly hastened and amplified in JOHD compared to AOHD [39]. Despite these initial observations from the Kids-JOHD study, larger-scale longitudinal studies in JOHD are still lacking. To this end, we have developed the first ever multi-site, prospective and comprehensive study of JOHD, leveraging the established infrastructure of the titular ChANGE-HD study to record clinical and neuroimaging metrics in this population. This study aims to investigate whether comprehensive longitudinal quantitative assessments of motor skills and cognition and neuroimaging in JOHD patients can yield reliable biomarkers of disease progression. A total of 40 patients with JOHD will be recruited. In the first year, 20 patients with JOHD (3-4 per site) will complete baseline assessments, with the remaining 20 patients undergoing baseline assessments in Year 2. Year 2 will also include annual follow-up visits for the first 20 patients. In Year 3, follow-up visits will be conducted for the second group of 20 patients. For the study, the comparator group will include GNE participants recruited in the ChANGE-HD protocol. To participate, JOHD participants must:

- Be diagnosed with JOHD by a trained neurologist prior to the age of 21.
- Be between 6-30 years old at baseline. Thus, even though the diagnosis is made prior to 21, assessment for the study is not restricted to ages under 21.
- Not be bed-ridden, unable to communicate, or beyond travel conditions that are too strenuous for the individual.

## DISCUSSION

ChANGE-HD is a longitudinal study designed to prospectively examine the neurodevelopmental phase of HD in premanifest at-risk children and young adults. Recruitment is currently underway at all six clinical sites. The data collected will provide crucial insights into the effects of *mHTT* and help uncouple neurodevelopmental changes from neurodegeneration.

Although recruitment is still ongoing, there are some prospects of how the results of ChANGE-HD may support the neurodevelopmental theory of HD. One such way is through antagonistic pleiotropy, as investigated by Neema et al. (2024) using the neuroimaging data of Kids-HD [17]. Therein, cortical hypertrophy and cognitive advantage were identified in the GE cohort very far from motor onset. If similar results are identified with the broader sample of ChANGE-HD, this would provide further support for the theory that *mHTT* confers developmental advantages antecedent to the degenerative phase.

Linked to this proposal is the theory of glutamate excitotoxicity in HD. It is a well-established fact that many neurological disorders are grounded in excessive neurotransmission through glutamatergic pathways which can then cause the development of epilepsy and/or neuronal atrophy [40–43]. Cortico-striatal pathways, which are highly implicated in the pathogenesis of HD, are primarily glutamatergic in nature [44]. If cortical hypertrophy is indeed characteristic of the developmental phase of HD, it is possible that a concomitant increase in glutamatergic neurotransmission is present as well. If this is true, there may be a direct causal link between developmental hypertrophy and downstream atrophy of striatal medium spiny neurons, which are the most affected neurons in HD. Neurochemical data will be needed to confirm this hypothesis, but nevertheless, the glutamatergic toxicity theory is a leading theory which connects two ostensibly dissonant phases of HD. The ChANGE-HD study is uniquely designed to test its validity.

## CONCLUSIONS

The ongoing ChANGE-HD study is the only multi-site longitudinal study in the world specifically designed to prospectively examine the neurodevelopmental effects of *mHTT* on children, adolescents, and young adults at risk for HD. While the study has broad applications, its primary objectives are to identify early neurodevelopmental changes, deepen our understanding of disease pathogenesis, and guide the design of therapeutic interventions. Together with the prospective JOHD study, ChANGE-HD will provide valuable insights into key aspects of disease progression in HD.

## DATA AVAILABILITY

No datasets were generated or analyzed during the current study. All relevant data from this study will be made available upon study completion.

## AUTHOR CONTRIBUTIONS

**Mohit Neema** – Data Curation; Formal Analysis; Investigation; Methodology; Software; Writing - Original Draft; Project Administration; Validation; Visualization; Writing – Review and Editing

**Nabil Nalabi** – Data Curation; Formal Analysis; Investigation; Methodology; Writing – Original Draft; Project Administration; Validation; Visualization; Writing – Review and Editing

**Peggy C. Nopoulos** – Conceptualization; Data Curation; Formal Analysis; Investigation; Methodology; Funding Acquisition; Project Administration; Resources; Supervision; Validation; Visualization; Writing – Review and Editing

## REFERENCES

1. MacDonald ME, Ambrose CM, Duao MP, Myers RH, Lin C, Srinidhi L, et al. A novel gene containing a trinucleotide repeat that is expanded and unstable on Huntington’s disease chromosomes. Cell. 1993;72: 971–983.

2. Cattaneo E, Zuccato C, Tartari M. Normal huntingtin function: An alternative approach to Huntington’s disease. Nature Reviews Neuroscience. 2005;6: 919–930.

3. Iennaco R, Formenti G, Trovesi C. The evolutionary history of the polyQ tract in huntingtin sheds light on its functional pro-neural activities. Cell Death and Differentiation. 2022;29: 293–305.

4. Hannan AJ. TRPing up the genome: Tandem repeat polymorphisms as dynamic sources of genetic variability in health and disease. Discovery Medicine. 2010;10: 314–321.

5. Frenkel ZM, Trifonov EN. Origin and evolution of genes and genomes. Crucial role of triplet expansions. Journal of Biomolecular Structure and Dynamics. 2012;30: 201–210.

6. Nopoulos PC. Huntington disease: A single-gene degenerative disorder of the striatum. Dialogues Clin Neurosci. 2016;18: 91–98.

7. van der Plas E, Schultz JL, Nopoulos PC. The Neurodevelopmental Hypothesis of Huntington’s Disease. Journal of Huntington’s Disease. 2020;9: 217–229.

8. The HD iPSC Consortium. Developmental alterations in Huntington’s disease neural cells and pharmacological rescue in cells and mice. Nature Neuroscience. 2017;20: 648–660.

9. Barnat M, Capizzi M, Aparicio E. Huntington’s disease alters human neurodevelopment. Science. 2020;369: 787–793.

10. Tabrizi SJ, Scahill RI, Owen G. Predictors of phenotypic progression and disease onset in premanifest and early-stage Huntington’s disease in the TRACK-HD study: Analysis of 36-month observational data. The Lancet Neurology. 2013;12: 637–649.

11. Rodrigues F, Byrne L, Tortelli R, Johnson EB, Wijeratne PA, Arridge M, et al. Mutant huntingtin and neurofilament light have distinct longitudinal dynamics in Huntington’s disease. Science Translational Medicine. 2020;12.

12. Long JD, Paulsen JS. Multivariate prediction of motor diagnosis in Huntington’s disease: 12 years of PREDICT-HD. Movement Disorders. 2015;30: 1664–1672.

13. Lee JK, Ding Y, Conrad AL, Cattaneo E, Epping E, Mathews K, et al. Sex-specific effects of the Huntington gene on normal neurodevelopment. Journal of Neuroscience Research. 2017;95: 398–408.

14. Schultz JL, Van Der Plas E, Langbehn DR, Conrad AL, Nopoulos PC. Age-related cognitive changes as a function of CAG repeat in child and adolescent carriers of mutating huntington. Annals of Neurology. 2021;89: 1036–1040. doi:10.1002/ana.26039

15. van der Plas E, Langbehn DR, Conrad AL, Koscik TR, Tereshchenko A, Epping EA, et al. Abnormal brain development in child and adolescent carriers of mutant huntingtin. Neurology. 2019;93: e1021–e1030.

16. Byrne LM, Schultz JL, van Der Plas, Langbehn D, Nopoulos PC, Wild EJ, et al. Neurofilament light protein as a potential blood biomarker for Huntington’s Disease in children. Movement Disorders. 2022;37: 1526–1531.

17. Neema M, Schultz JL, Langbehn DR, Conrad AL, Epping EA, Magnotta VA, et al. Mutant huntingtin drives development of an advantageous brain early in life: Evidence in support of antagonistic pleiotropy. Annals of Neurology. 2024;96: 1006–1019.

18. Williams GC. Pleiostropy, natural selection and the evolution of senescence. Evolution. 1957;11: 398–411.

19. Lebel C, Beaulieu C. Longitudinal development of human brain wiring continues from childhood into adulthood. Journal of Neuroscience. 2011;31: 10937–10947.

20. Weintraub S, Dikmen SS, Heaton RK, Tulsky DS, Zelazo PD, Bauer PJ, et al. Cognition assessment using the NIH Toolbox. Neurology. 2013;80:S54–64: S54–S64.

21. van der Plas E, Schubert R, Reilmann R, Nopolous PC. A feasibility study of quantitative motor assessments in children using the q-motor suite. Journal of Huntington’s Disease. 2019;8: 333–338.

22. Iverson GL, Connors EJ, Marsh J, Terry DP. Examining normative reference values and item-level symptom endorsement for the quality of life in neurological disorders (Neuro-QoL) v2.0 cognitive function-short form. Archives of Clinical Neuropsychology. 2021;36: 126–134.

23. Bruni O, Ottaviano S, Guidetti V, Romoli M, Innocenzi M, Cortesi F, et al. The sleep disturbance scale for children (SDSC). Construction and validation of an instrument to evaluate sleep disturbances in childhood and adolescence. Journal of Sleep Research. 1996;5: 251–261.

24. Casey BJ, Cannonier T, Conley MI, Cohen AO, Barch DM, Heitzeg MM, et al. The adolescent brain cognitive development (ABCD) study: Imaging acquisition across 21 sites. Developmental Cognitive Neuroscience. 2018;32: 43–54.

25. Coupe P, Mansencal B, Clement M, Giraud R, Denis de Senneville B, Ta V-T, et al. AssemblyNet: A large ensemble of CNNs for 3D whole brain MRI segmentation. NeuroImage. 219: 117026.

26. Fischl B. FreeSurfer. NeuroImage. 2012;62: 774–781. doi:10.1016/j.neuroimage.2012.01.021

27. Cieslak M, Cook PA, He X, Yeh F-C, Dhollander T, Adebimpe A, et al. QSIPrep:Aan integrative platform for preprocessing and reconstructing diffusion MRI data. Nature Methods. 2021;18: 775–778. doi:10.1038/s41592-021-01185-5

28. Damoiseaux JS, Rombouts SARB, Barkhof F, Scheltens P, Stam CJ, Smith SM, et al. Consistent resting-state networks across healthy subjects. Proc Natl Acad Sci USA. 2006;103: 13848–13853. doi:10.1073/pnas.0601417103

29. Finn ES, Shen X, Scheinost D, Rosenberg MD, Huang J, Chun MM, et al. Functional connectome fingerprinting: Identifying individuals using patterns of brain connectivity. Nat Neurosci. 2015;18: 1664–1671. doi:10.1038/nn.4135

30. Esteban O, Markiewicz CJ, Blair RW, Moodie CA, Isik AI, Erramuzpe A, et al. fMRIPrep: a robust preprocessing pipeline for functional MRI. Nat Methods. 2019;16: 111–116. doi:10.1038/s41592-018-0235-4

31. Mehta K, Salo T, Madison TJ, Adebimpe A, Bassett DS, Bertolero M, et al. XCP-D: A robust pipeline for the post-processing of fMRI data. Imaging Neuroscience. 2024;2: imag–2–00257. doi:10.1162/imag_a_00257

32. Vijayakumar N, Mills KL, Alexander-Bloch A, Tamnes CK, Whittle S. Structural brain development: A review of methodological approaches and best practices. Developmental Cognitive Neuroscience. 2018;33: 129–148.

33. Langbehn DR, Brinkman RR, Falush D, Paulsen JS, Hayden MR. A new model for prediction of the age of onset and penetrance for Huntington’s disease based on CAG length. Clinical Genetics. 2004;65: 267–277.

34. Oosterloo M, Touze A, Byrne LM, Achenbach J, Aksoy H, Coleman A, et al. Clinical review of Juvenile Huntington’s Disease. Journal of Huntington’s Disease. 2024;13: 149–161.

35. Stout JC. Juvenile Huntington’s Disease: Left behind? Lancet Neurol 2018;17:932–933. The Lancet Neurology. 17: P932-933.

36. Tereshchenko A, McHugh M, Lee JK, Gonzalez-Alegre P, Crane K, Dawson J, et al. Abnormal weight and body mass index in children with Juvenile Huntington’s Disease. Journal of Huntington’s Disease. 2015;4: 231–238.

37. Tereshchenko A, Plas E, Mathews KD. Developmental Trajectory of Height, Weight, and BMI in Children and Adolescents at Risk for Huntington’s Disease. Effect of mHTT on Growth J Huntingtons Dis. 2020;9: 245–251.

38. Tereshchenko A, Magnotta VA, Epping E, Mathews K, Espe-Pfeifer, Martin E, et al. Brain structure in juvenile-onset Huntington disease. Neurology. 2019;92: e1939–e1947.

39. Schultz JL, Langbehn DR, Al-Kaylani HM, van Der Plas E, Koscik TR, Epping EA, et al. Longitudinal clinical and biological characteristics in Juvenile-Onset Huntington’s Disease. Movement Disorders. 2023;38: 113–122.

40. Salinska E, Danysz W, Lazarewicz JW. The role of excitotoxicity in neurodegeneration. Folia Neuropatholica. 2005;43: 322–339.

41. Mehta A, Prabhakar M, Kumar P, Deshmukh R, Sharma PL. Excitotoxicity: Bridge to various triggers in neurodegenerative disorders. European Journal of Pharmacology. 2013;698: 6–18.

42. Dong X, Wan Y, Qin Z. Molecular mechanisms of excitotoxicity and their relevance to pathogenesis of neurodegenerative diseases. Acta Pharmacologica Sinica. 2009;30: 379–387.

43. Doble A. The role of excitotoxicity in neurodegenerative disease: Implications for therapy. Pharmacology & Therapeutics. 1999;81: 163–221.

44. Haber SN. Corticostriatal circuitry. Dialogues in Clinical Neuroscience. 2016;18: 7–21.

